# Clinical Presentation, Renal Histopathological Findings and Outcome in Patients with Monoclonal Gammopathy and Kidney Disease

**DOI:** 10.1101/2020.06.21.20136739

**Authors:** Gaetano Alfano, Alice Delrio, Francesco Fontana, Giacomo Mori, Annachiara Ferrari, Rossella Perrone, Silvia Giovanella, Giulia Ligabue, Riccardo Magistroni, Gianni Cappelli

## Abstract

Monoclonal gammopathies have been associated with kidney injury. Nephrotoxicity of the secreted monoclonal (M)-protein relies on a complex interplay between biological characteristics and serum concentration. Little is known about the epidemiology and clinical manifestations of the different types of monoclonal gammopathies in patients with kidney disease.

We enrolled all patients with monoclonal gammopathy who underwent kidney biopsy between January 2000 and March 2017. Data about demographics, clinical manifestations and histological lesions were collected retrospectively.

Monoclonal gammopathy was detected in 174 (13%) patients with a mean age of 66.4±13.1 years. M-protein was secreted by monoclonal gammopathy of undetermined significate (MGUS) (52,8%), myeloma multiple (MM) (25.2%), primary amyloidosis (AL) (9,1%), smoldering MM (7 %), non-Hodgkin lymphoma (NHL) (6.8%) and HL (1.7%). Monoclonal gammopathy of renal significance (MGRS) accounted for 6.5% in patients with MGUS and 14.2% in patients with smoldering MM. Evaluation of kidney biopsy revealed that M-protein was directed involved in causing kidney injury in MM (93.1%) and NHL (8,3%). MM was the only gammopathy significantly associated with an increased risk of kidney injury (odds ratio [OR]=47.5, CI95%, 13.7-164.9; P=<0.001). While there were no significant differences in the progression toward end-stage renal disease or dialysis (P=0.776), these disorders were associated with a different risk of death (P=0.047) at the end of the follow-up.

Monoclonal gammopathy was a frequent finding in patients with kidney disease. Kidney biopsy had a key role in identifying the underlying monoclonal gammopathy and recognizing the causal relationship between M-protein and kidney injury.

## Introduction

Monoclonal gammopathy is a clinical condition characterized by the presence of an abnormal protein — known as monoclonal (M)-protein or paraprotein— in the blood.[1] M-protein is an intact antibody, or any chain fragment produced and secreted by a pathological lymphoproliferative clone. The ability of M-protein to disrupt cellular homeostasis is unpredictable and principally related to its physicochemical properties and blood concentration.[2]

Historically, renal toxicity of the M-protein has been associated with the malignancy of the underlying lymphoproliferative disorder. Multiple Myeloma (MM), one of the most common hematologic malignancy[3], has been widely associated with kidney disease.[4,5]. Renal injury in this disease relies principally on the overproduction of M-protein, which, freely filtered by the glomerulus rapidly overwhelms tubular reabsorption capacity, leading to intraluminal precipitation with tubular obstruction and activation of inflammatory pathways. As a result, cast nephropathy is the classical renal presentation of MM[6]. Other malignant lymphoproliferative diseases, such as Waldenström macroglobulinemia(WM)[7], lymphomas[8,9], have been frequently associated with renal injury and can present with a wide range of glomerular injury on kidney biopsy including M-immunoglobulin deposition disease, proliferative glomerulonephritis with M-immunoglobulin deposit and cryoglobulinemic vasculitis.

Even clones that have a low propensity to progress toward malignancy and secrete a low amount of M-protein can be involved in tissue damage including neuropathy, autoimmune diseases as well as kidney disease.[10] In this setting, the deposition of M-protein (direct mechanism) or activation of the complement system without tissue deposition of M-immunoglobulin (indirect mechanism) may lead to renal lesions[11]. Recently, the nephrological community has coined a new term: monoclonal gammopathy of renal significance (MGRS). The definition of MGRS includes all small B-cell or plasma cell clones that, per se, do not meet strict hematological criteria for cytoreductive therapy but are implied in kidney injury through the production of M-protein.[12] The most glaring example is monoclonal gammopathy of undetermined significate (MGUS), which may be associated with renal lesions despite the low level of secreted paraprotein and the rare progression to MM.[13] The growing interest in this new pathological entity has spurred nephrologists and hematologists to reevaluate the pathogenicity of these small, apparently indolent, clones. The enthusiasm to understand the causative role of M-protein in patients with renal impairment was tempered by the rarity of the phenomena. Considering these limits and the fragmentary of the data published in the literature, we explored the association between monoclonal gammopathies and kidney disease with the aim to define prevalence, clinical manifestations as well as outcomes of these disorders in a cohort of patients who underwent kidney biopsy for renal impairment.

## Material and Methods

We conducted a retrospective study at the Nephrology Unit of the University Hospital of Modena. The medical charts of all patients with biopsy-proven kidney disease were evaluated from January 2000 to March 2017.

Among all patients who underwent kidney biopsy, we enrolled only those with a diagnosis of serum M-protein that was detected by protein electrophoresis (SPEP) and subsequently characterized by serum or urine immunofixation. Hence, patients with a not-confirmed diagnosis of monoclonal gammopathies were excluded from the study.

The study protocol was approved by the Provincial Ethics Committee of the University Hospital of Modena (CE/1476).

### Renal biopsy

Biopsy specimens were examined using light microscopy (LM) and immunofluorescence (IF). Kidney tissue sections have been evaluated by hematoxylin and eosin, periodic acid-Schiff (PAS), periodic acid-methenamine silver (Jones), and Masson’s trichrome stains. For immunofluorescence, cryostat sections were stained with fluorescein isothiocyanate-conjugated rabbit antihuman IgG, IgM, IgA, C3, C1q, kappa (k) and lambda (λ) light chain. Staining with Congo red was used to confirm amyloid deposits. Primary amyloidosis or light chain amyloidosis (AL) was identified by light chain restriction using IF or immunohistochemistry performed in another Center. Electron microscopy examination was performed only on pathologists’ indication.

Monoclonal gammopathy was considered directly involved in the pathogenesis of renal lesions if histological examination revealed immunoglobulin-associated kidney lesions with heavy or light chain restriction on IF (restriction for the same immunoglobulin isotype detected in blood). Diagnosis of cast nephropathy, M-immunoglobulin deposition disease, membranoproliferative glomerulonephritis with M-immunoglobulin deposit and primary amyloidosis or light chain amyloidosis (AL) were the classical kidney diseases directly associated with monoclonal gammopathy[11].

### Data collection and definition

Demographics and laboratory data were collected at the time of kidney biopsy. Data about complete blood count (leukocytes, erythrocytes, hemoglobin, platelets), serum calcium, sCr, estimated glomerular filtration(eGFR) calculated using CKD-EPI equation[14], proteinuria, serum and urine M-protein, serum M-protein concentration and serum-free light chain (FLC) were extracted from medical records.

Proteinuria was principally estimated through the urine protein-to-creatinine ratio.

Nephrotic syndrome was defined as urine protein-to-creatinine ratio more 3 mg/mg and serum albumin less than 3.5 gr/dl. We considered AL amyloidosis and light chain deposition disease associated with MM in the presence of lytic bone lesions and other sign of over MM or plasma cells count greater than 30% in the bone marrow.[15]

Acute kidney injury (AKI) refers to an abrupt decrease in kidney function. The definition is based on the following criteria: increase in serum creatinine (sCr) by ≥ 0.3 mg/dL within 48 hours, or increase in sCr level to ≥ 1.5 times from baseline.[16] The urinary criterium was not used for the diagnosis of AKI. Baseline creatine corresponded to the last sCr before kidney biopsy when it was not available, we considered the sCr measured at hospital admission.

Severe impairment of renal function refers to acute worsening of renal function with a serum sCr level ≥ 3 mg/dL.

After identification and characterization of the serum M-immunoglobulin, radiologic workup, flow cytometry or bone marrow biopsy were performed according to hematologist’s indications.

### Statistical analysis

Continuous variables with normal distribution were presented as mean and standard deviation (SD). The difference between the means of two groups was performed with a two-sample t-test or Mann Whitney test. One one-way ANOVA and Kruskal-Wallis test (or one-way ANOVA on ranks) were used to perform multiple comparisons between groups. Turkey’s test was used to determine the statistical difference between the mean of all possible pairs. Logistic regression analyses were done to compute odds ratios (AOR), 95% confidence intervals (95% CI), and P-values of lymphoproliferative disorders to predict direct kidney injury. Logistic regression was also used to evaluate the association between histological findings in MM patients and severe renal impairment. P-value <0.05 was considered to be statistically significant. All analyses were performed using SPSS version 23(SPSS, Inc., Chicago, IL).

## Results

### Patient’s characteristics

We reviewed the charts of 1334 patients who underwent native kidney biopsy to investigate renal dysfunction. Monoclonal gammopathy was found in 174 patients (13%) with a mean age of 66.4±13.1 years. Most of them were of Caucasian origin (96%), and males were predominant on females (67.2 vs. 32.8%) (Table 1). Compared to the entire study population, monoclonal gammopathy was more frequent in patients aged 50-79 years (Table 2).

**Table 1.** Demographics and clinical characteristics of patients with monoclonal gammopathy

| Characteristic | All patients |
| --- | --- |
| Age – yr |  |
| Mean (±SD) | 66.4±13.1 |
| Male – n. (%) | 112 (67.2%) |
| Follow-up – yr | 5.3–4.5 |
| Ethnic group – n. (%) |  |
| Caucasian | 167 (96) |
| Hispanica | 1 (0.6) |
| Asiatic | 3 (1.7) |
| African | 3 (1.7) |
| Monoclonal gammopathy |  |
| MGUS – n. (%) | 92 (52.8) |
| MM – n. (%) | 44 (25.2) |
| AL amyloidosis – n. (%) | 16 (9.1) |
| NHL – n. (%) | 12 (6.8) |
| Smoldering MM – n. (%) | 7 (4) |
| HL – n. (%) | 3 (1.7) |

HL denotes Hodgkin lymphoma, NHL, non-Hodgkin lymphoma; MGUS, monoclonal gammopathy of indeterminate significance; MM, myeloma multiple; SD, standard deviation; SMM, smoldering myeloma multiple.

**Table 2.**
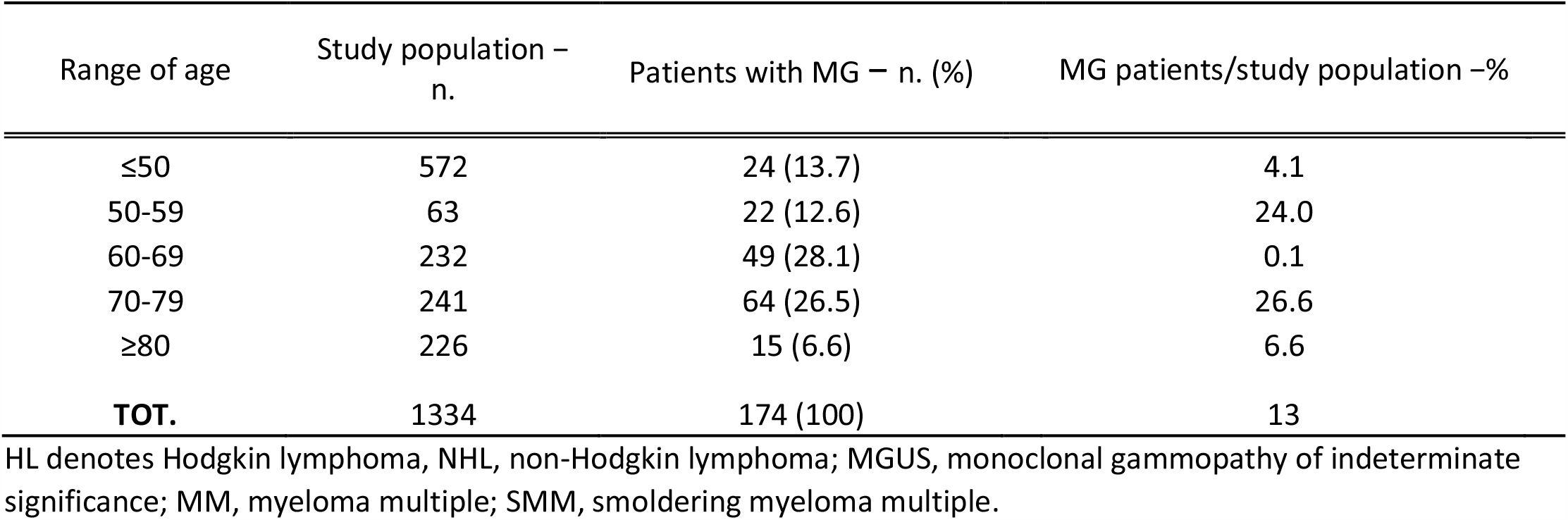
Range of age in patients with monoclonal gammopathy

The hematologic disorders presenting with M-protein in peripheral blood were MGUS (52.8%), MM (25.2%), amyloidosis (9.1%) smoldering MM (4%), non-Hodgkin lymphoma (NHL) (6.8%) and HL (1.7%)(Table 1 and Fig.1). The detection of M-protein was associated with 56.8% of pre-malignant lymphoproliferative diseases and with 43.2% of malignant diseases. There were no differences (P=0.16) between the age of patients with benign (5.3±13.7 years) and malignant lymphoproliferative disease (67.5±11.9 years).

**Figure 1.**
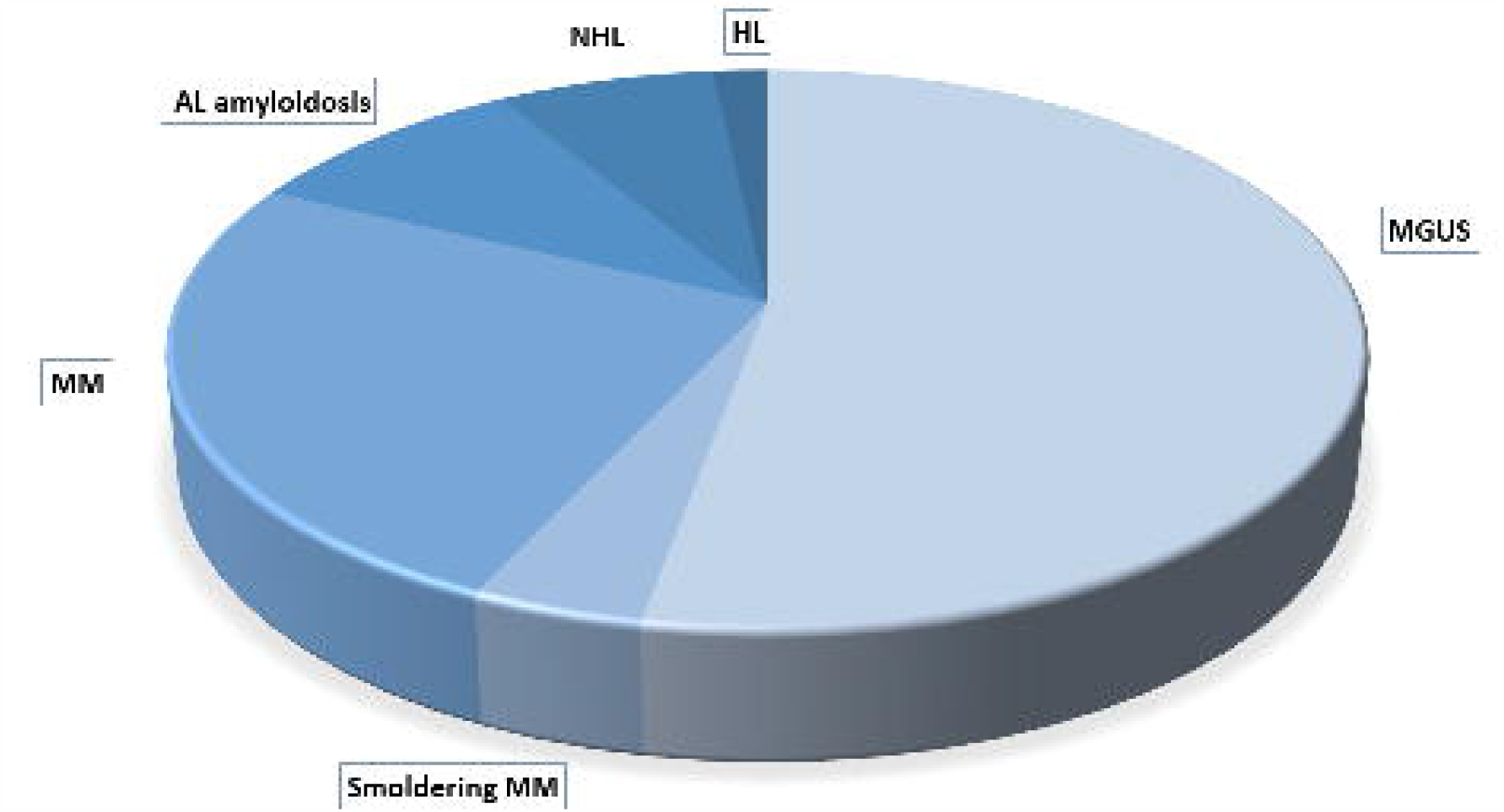
Distribution of monoclonal gammopathy in our cohort of patients

### MGUS

MGUS was the most common monoclonal gammopathy. Its prevalence was estimated to be 52.8% among patients with serum M-protein.

The mean age of patients was 64.9±13.9 years with a predominance of males 62% (Table3). Average sCr was 2.68±2.1 mg/dl corresponding to an estimated glomerular filtration rate (eGFR) of 35.2±29.3 ml/min. Urinary examination showed a urine protein-to-creatinine ratio of 5.1±6.5.

The serum concentration of M-protein oscillated between 0.2 and 2.3 gr/dl with a mean concentration of 0.6±0.5 gr/dl. IgG was the most common isotype (65.5%), followed by IgM (24.7%) and IgA (2.15%) Characterization of serum M-protein through immunofixation revealed a higher prevalence of *k* light chain; M-protein was biclonal in 7.5% of the cases.

Serum FLC *k* and λ were 302.3±177.8 and 172.5±159.3, respectively. Bence Jones proteinuria was detected in 28.2% of the subjects.

In 16 (17.3%) patients, the histological evaluation revealed glomerular lesions compatible with a membranoproliferative glomerulonephritis pattern. It was secondary to viral hepatitis in 4 subjects. Further kidney diseases included membranous glomerulopathy (16.3%), end-stage kidney disease (16.3%), ANCA-associated vasculitis (8.6%), postinfectious glomerulonephritis (7.6%), focal segmental glomerular sclerosis (5.4); and interstitial nephritis (4.3%). Surprisingly, IgA glomerulonephritis was underrepresented in this group (1%).

In four patients (4.3%) MGUS was directly involved in the development of kidney injury through the deposition of light chains, and in two cases with a diagnosis of kidney disease with pattern of glomerular injury, IF analysis showed intact immunoglobulin restriction (lesions compatible with proliferative glomerulonephritis with Ig deposit. Overall, parenchymal lesions compatible with MGRS criteria accounted for 6.5% of all cases of MGUS.

At the end of the observational period of 5.4 years, 37% of patients died (Table 5) and 32% were on replacement renal therapy (Table 6).

**Table 3.**
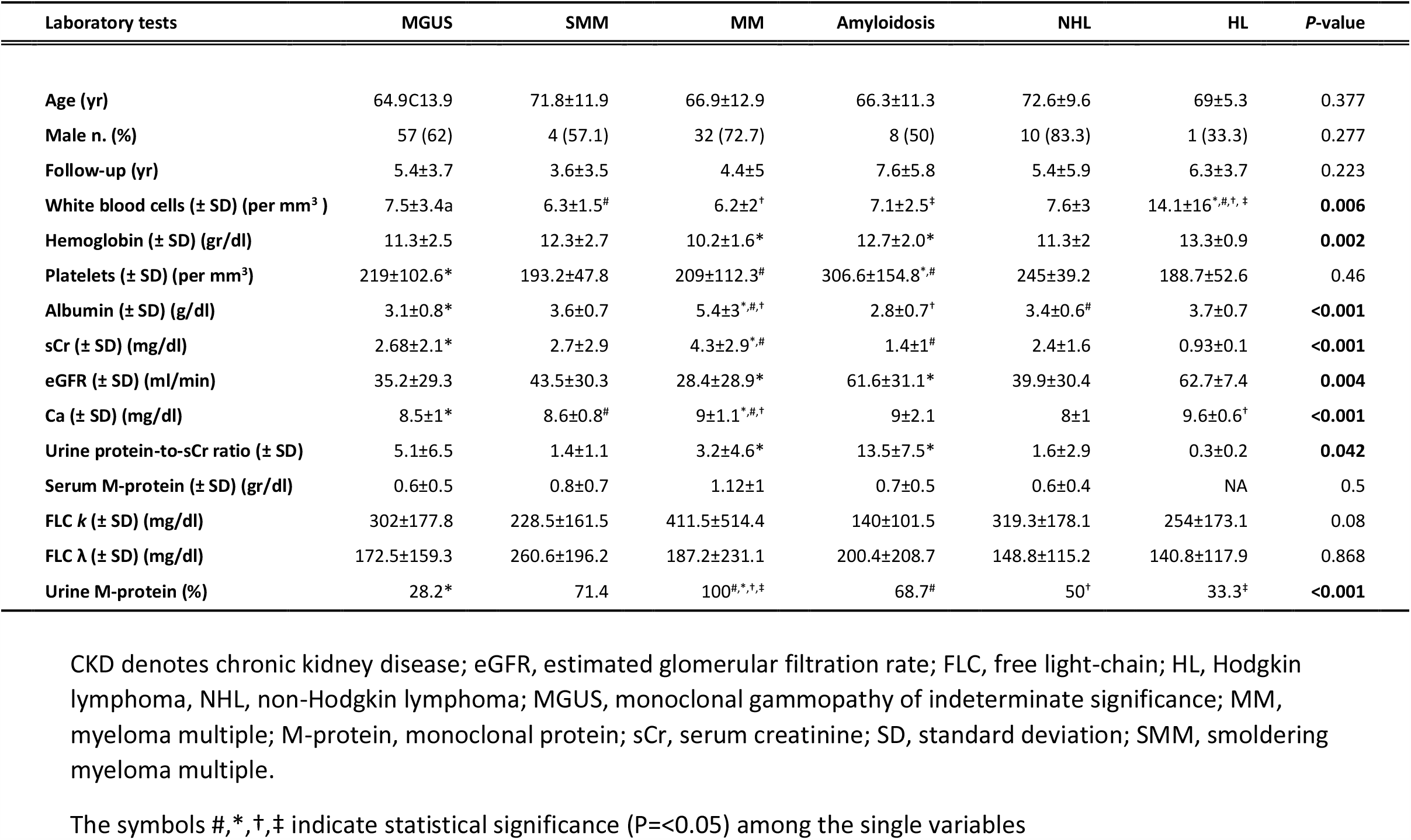
Clinical characteristics and lab examinations of patients with monoclonal gammopathy

**Table 4.** Association between monoclonal gammopathies and direct kidney injury

| Monoclonal gammopathy | Odds ratio (95% CI)* | P-value |
| --- | --- | --- |
| MM | 47.5 (13.7-164.9) | **<0.001** |
| NHL | 2.1 (0.6-7.2) | 0.194 |
| SMM | 0.5 (0.1-3) | 0.5 |
| MGUS | 0.01 (0.005-0.04) | **<0.001** |
| HL | / | 0.153 |

HL denotes Hodgkin lymphoma, NHL, non-Hodgkin lymphoma; MGUS, monoclonal gammopathy of indeterminate significance; MM, myeloma multiple; SMM, smoldering myeloma multiple.
AL amyloidosis was not tested because the pathogenesis of amyloidosis depends on a proven direct kidney injury due to the deposition of light-chain within the renal parenchyma.
P-value for MGUS was statistically but not clinically significant. The protective effect of MGUS in developing kidney disease is not applicable in this context.

**Table 5.**
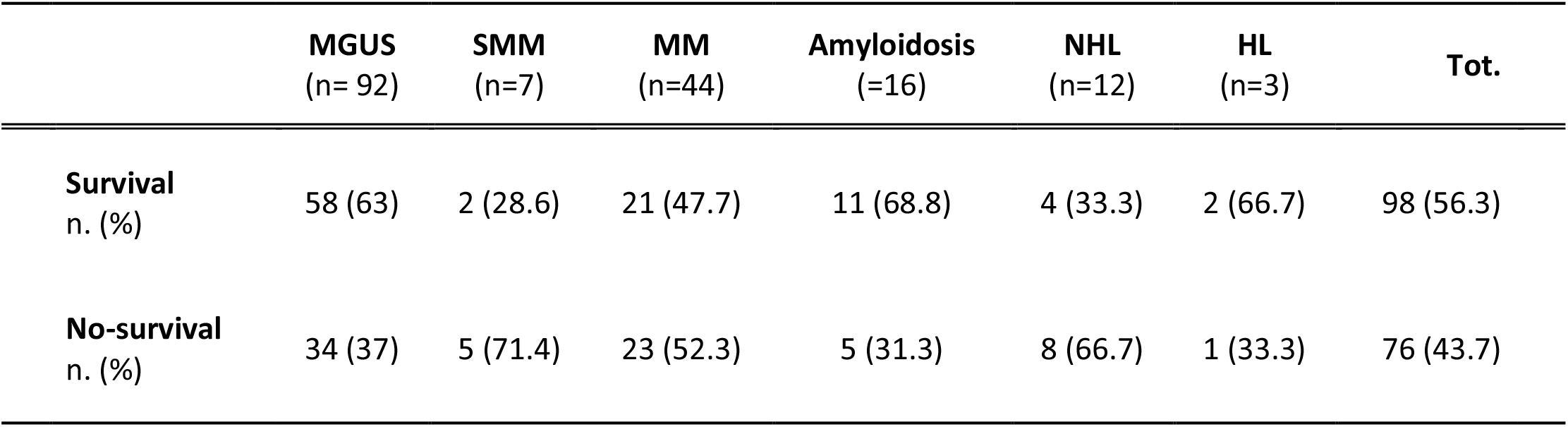
Case fatality rate in patients with monoclonal gammopathy

|  | **MGUS**<br>(n= 92) | **SMM**<br>(n=7) | **MM**<br>(n=44) | **Amyloidosis**<br>(=16) | **NHL**<br>(n=12) | **HL**<br>(n=3) | **Tot.** |
| --- | --- | --- | --- | --- | --- | --- | --- |
| **Survival**<br>n. (%) | 58 (63) | 2 (28.6) | 21 (47.7) | 11 (68.8) | 4 (33.3) | 2 (66.7) | 98 (56.3) |
| **No-survival**<br>n. (%) | 34 (37) | 5 (71.4) | 23 (52.3) | 5 (31.3) | 8 (66.7) | 1 (33.3) | 76 (43.7) |

**Table 6.** Rate of end-stage renal disease or dialysis in patients with monoclonal gammopathy

|  | **MGUS**<br>(n= 92) | **SMM**<br>(n=7) | **MM**<br>(n=44) | **Amyloidosis**<br>(=16) | **NHL**<br>(n=12) | **HL**<br>(n=3) | **Tot.\*** |
| --- | --- | --- | --- | --- | --- | --- | --- |
| **CKD stage 1-4**<br>n. (%) | 57 (67.9) | 5 (83.3) | 21 (61.8) | 11 (68.8) | 6 (66.7) | 3 (100) | 103 (67.8) |
| **CKD stage 5/Dialysis**<br>n. (%) | 27 (32.1) | 1 (16.7) | 13 (38.2) | 5 (31.3) | 3 (33.3) | 0 (3) | 49 (32.2) |

\*Missing data =13%
CKD denotes chronic kidney disease; HL, Hodgkin lymphoma, NHL, non-Hodgkin lymphoma; MGUS, monoclonal gammopathy of indeterminate significance; MM, myeloma multiple; SMM, smoldering myeloma multiple.

### Multiple Myeloma

MM was the second most common gammopathy in our study population. The disorder was detected in 44 subjects with a mean age of 66.9±12.9 years. MM was more frequent in males than in females (72.7 vs 27.3%). According to the criteria CRAB (hypercalcemia, renal disease, anemia and bone disease), laboratory tests at presentation revealed serum calcium of 9±1.1 mg/dl, hemoglobin of 10.2±1.6 gr/dl, myeloma bone lesions in 75% of patients and average sCr of 4.3±2.9 mg/dl, corresponding to an eGFR of 28.4.7±28.9 ml/min (Table3). Plasma cells infiltrate in bone marrow biopsy accounted for 50% of the cells. The majority of the patients (81.8%) was admitted with severe impairment of renal function and 27.2% needed urgent kidney replacement therapy. In seven patients (15.9%) clinical manifestation of kidney disease was nephrotic syndrome with average urine protein-to-creatinine ratio ranging from 4.2 to 18.5 associated with a wide variability of renal function (sCr ranged from 1.01 to 6 mg/dl).

Light chain MM accounted for 34.1% of the cases and, as expected, kappa light chain MM resulted more prevalent compared to λ light-chain MM (60 vs. 40%). M-protein isotype were IgGk (22.7%); IgGλ (15.9%); IgAk (6.8%); IgA λ (13.6%) and IgMk (6.8%). Bence Jones was detected in 93.1% of the tested patients.

Histological evaluation of kidney biopsy specimens showed cast nephropathy (68.1%), AL amyloidosis (15.9%), light chain deposition disease (6.8%) and interstitial nephritis (9.1%). Cast nephropathy was the only histological lesion associated with severe renal impairment (OR=26.2, 95%CI, 2.8-245.5; P=0.004) (supplemental Table1). Lastly, all patients with histological diagnosis different from cast nephropathy had bone osteolytic lesions compatible with myeloma bone disease. The case fatality rate was high (52.3%) and more than one-third of the patients was on ESRD (38.2%) at the end of the study (Table 5 and 6).

### Smoldering MM

Seven (4%) patients had a diagnosis of smoldering MM. The disorder manifested at a mean age of 71.8±11.9 years and showed a slightly higher prevalence in men than women (72.7 vs 27.3%).

According to the definition of MM smoldering, bone lytic lesions were absent in all patients. Hemoglobin and serum calcium were in the normal range, 12.3±2.3 gr/dl and 8.6±0.8 mg/dl, respectively (Table 3). At presentation, mean sCr was 2.7±2.9 mg/dl (eGFR of 43.5±30.3ml/min) with proteinuria of 1.4±1.1 mg/mg. Immunofixation of the serum M-protein detected the following isotypes: IgGλ (28.2%), IgGk (28.2%)IgMk (14.1%), IgAλ (14.1%) and *k* light chain (14.1%). Bence Jones was found in 71.4% of the patients. Bone marrow biopsy revealed a mean plasma cell count of 18%.

Evaluation of renal biopsies showed different patterns of glomerular diseases including membranoproliferative glomerulonephritis (28.5%), interstitial nephritis (14.2%), light chain deposition disease (14.2%) acute tubular necrosis (ATN) (14.2%), ANCA-negative vasculitis (14.2%) and membranous glomerulonephritis (14.2%). Light chain restriction was diagnosed only in one patient (14.2%) affected by membranoproliferative glomerulonephritis.

### Hodgkin Lymphoma

Three patients (1.12%) had a diagnosis of Hodgkin’s lymphoma at an average age of 69.04±5.3 years. All patients had a normal renal function manifesting with a mean sCr of 0.93±0.07 mg/dl corresponding to an eGFR of 62.7.3±7.4 ml/min. Mean urine protein-to-creatinine ratio was of 0.3±0.2 (Table 3). Mild proteinuria was present only in one patient (urine protein-to-creatinine ratio of 0.5). Serum immunofixation identified isotype IgMk in two cases and isotype IgGk in only one case. Mean FLC k was and free light chain lambda were 254±173.1 and 140.8±117.9 mg/dl, respectively. Bence Jones protein was present in only one patient. Cryoglobulinemic glomerulonephritis was found in two-thirds of the patients (66.6%) and hypertensive nephrosclerosis in one (33.3%).

### AL Amyloidosis

Amyloidosis, defined as primary amyloidosis or AL amyloidosis AL was diagnosed in 16 patients (9.1%). Amyloidosis was secondary to MGUS (75%) and smoldering MM (25%). The mean age of the affected subjects was 66.34 ±11.38 years and females were slightly more prevalent than males (53 vs 46%). sCr ranged from 0.5 to 4.5 mg/dl with a mean level of 1.4 gr/dl, corresponding to 56.5 ml/min of eGFR. Nephrotic syndrome was the most common presentation (75%). Overall, patients presented with high proteinuria (8.33±3.2) associated to hypoalbuminemia (2.74±0.84 gr/dl) (Table 3).

Serum immunofixation revealed the following isotypes: IgGλ (56%), IgGk (13%), IgAλ (13%), IgM*k* (13%) and IgMλ (6%). Serum M-protein concentration was 0.7±0.6 gr/dl. Detention of urinary light chain occurred in 68.7% of the patients.

The diagnosis of amyloidosis was performed by the detection of deposit of amorphous material in the mesangium and capillary loops of glomeruli. Congo red stain confirmed the diagnosis of amyloidosis and immunohistochemical analysis identified on kidney tissue the corresponding serum light chain as the cause of amyloid deposition.

### Non-Hodgkin’s lymphoma

Twelve patients (6.8%) with monoclonal gammopathy had a diagnosis of NHL, which term includes several heterogeneous lymphoproliferative disorders. According to the WHO classification[17] lymphoplasmacytic lymphoma accounted for 41.6%, Waldenstrom’s macroglobulinemia for 30.7%, marginal zone lymphoma for 15.2%, diffuse large B-cell lymphoma for 7.6% and anaplastic large cell lymphoma for 7.6%. Male gender was fully associated (100%) with NHL in our cohort of patients. The average age of subjects was 72.6±9.6 years. Renal function was extremely variable at presentation, with sCr ranging between 0.85 and 6.35 mg/dl; mean sCr was 2.4±1.6 mg/dl corresponding at an eGFR of 30.4±22.7 ml/min. Five out of 12 patients had nephrotic syndrome at hospital-admission. Daily proteinuria ranged from 0.7 to 8.2 gr/day. Average proteinuria was 4.36±3.36 gr/24 hour IgM*k* (66.6%) was the most common monoclonal protein whereas IgA lambda, IgG *k*, IgM λ accounted for 8.33%, respectively. A patient with marginal zone lymphoma had a circulating biclonal M-protein NHL subjects had a circulating M-protein of 0.6±0.4 gr/dl. Urine monoclonal component was found in 68.7% of patients

Histological evaluation of biopsy specimen revealed amyloidosis (25%), glomerulonephritis pattern of injury (16.6%), LCDD (25%), ANCA-associated vasculitis (8.3%), cast-nephropathy (8.3%), and hypertensive nephrosclerosis (8.3%). In one case (8.3%) Ig deposits were restricted for the same serum M-protein in a context of a membranoproliferative pattern of glomerular injury.

### Comparison between groups

Kruskal-Wallis test showed that mean serum sCr levels (P= < 0.0001), eGFR (P=0.004), proteinuria (P= < 0.042), serum calcium(P= < 0.0001), serum albumin(P= < 0.0001), white blood count(P= < 0.006) and hemoglobin (P= < 0.002) were statistically different between the groups (Table 3).

Lymphoproliferative diseases were variably associated with renal lesions due to M-protein. Documented direct kidney injury in patients with NHL, MGUS, MM, SMM and AL amyloidosis accounted for 58.3%,6.5%, 91.3%,14.1% and 100%, respectively. Regression analysis excluding amyloidosis, which diagnosis necessarily required demonstration of tissue damage, showed that MM was significantly associated with renal damage (P= <0.001). MM patients had a 47.5-fold increased risk of renal lesions (95 %CI, 13.7-164.9).

There were no statistically significant differences in crude case-rate fatality (P=0.113) and incidence of ESRD or dialysis (P= 0.751) among groups with monoclonal gammopathies. Kaplan Meier survival analysis revealed a small difference in the survival of patients with monoclonal gammopathy (P=0.047) (Fig.2) and confirmed that there were no differences in the incidence of ESRD or dialysis (Fig.3).

**Figure 2.**
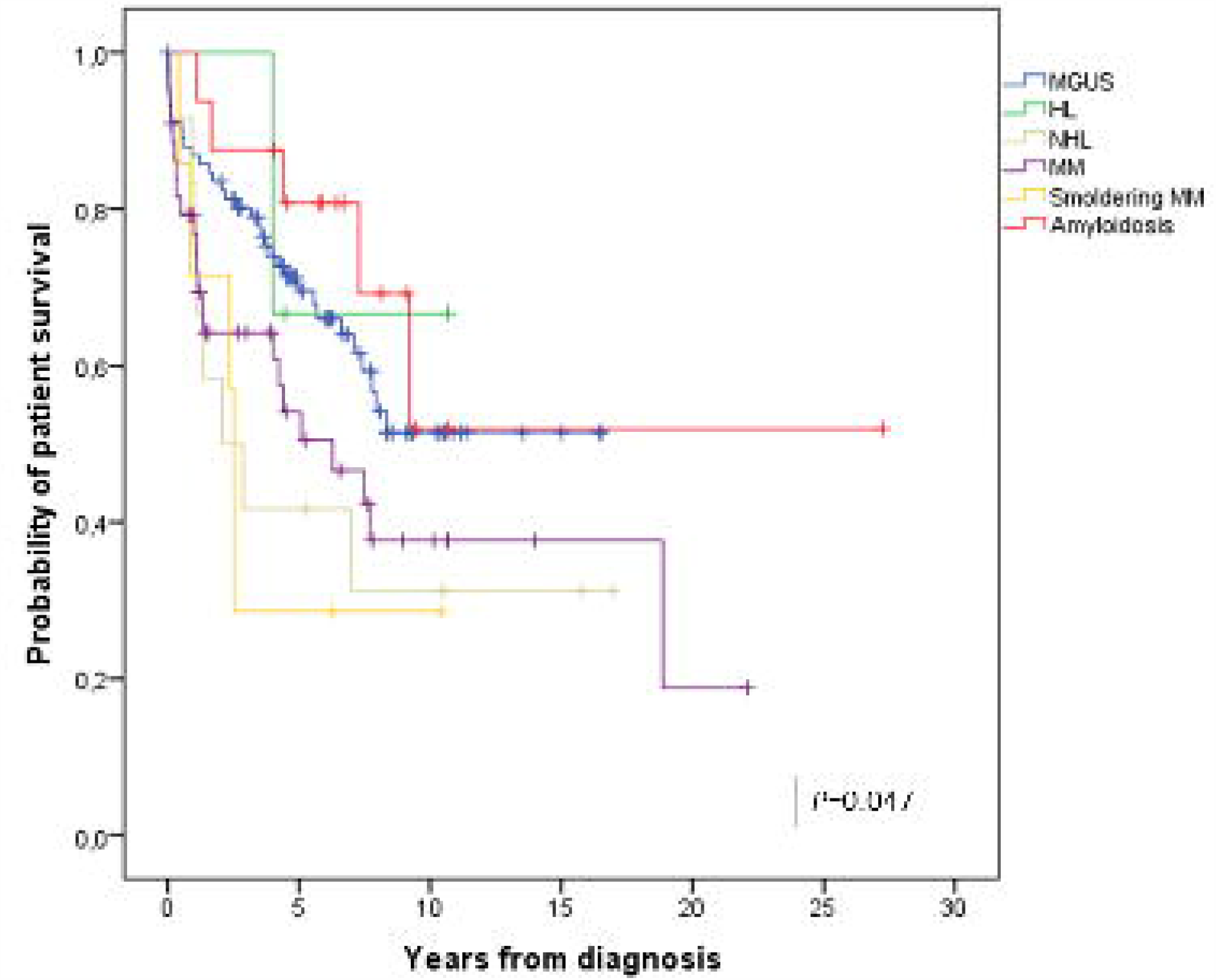
Overall survival according to the diagnosis of monoclonal gammopathy

**Figure 3.**
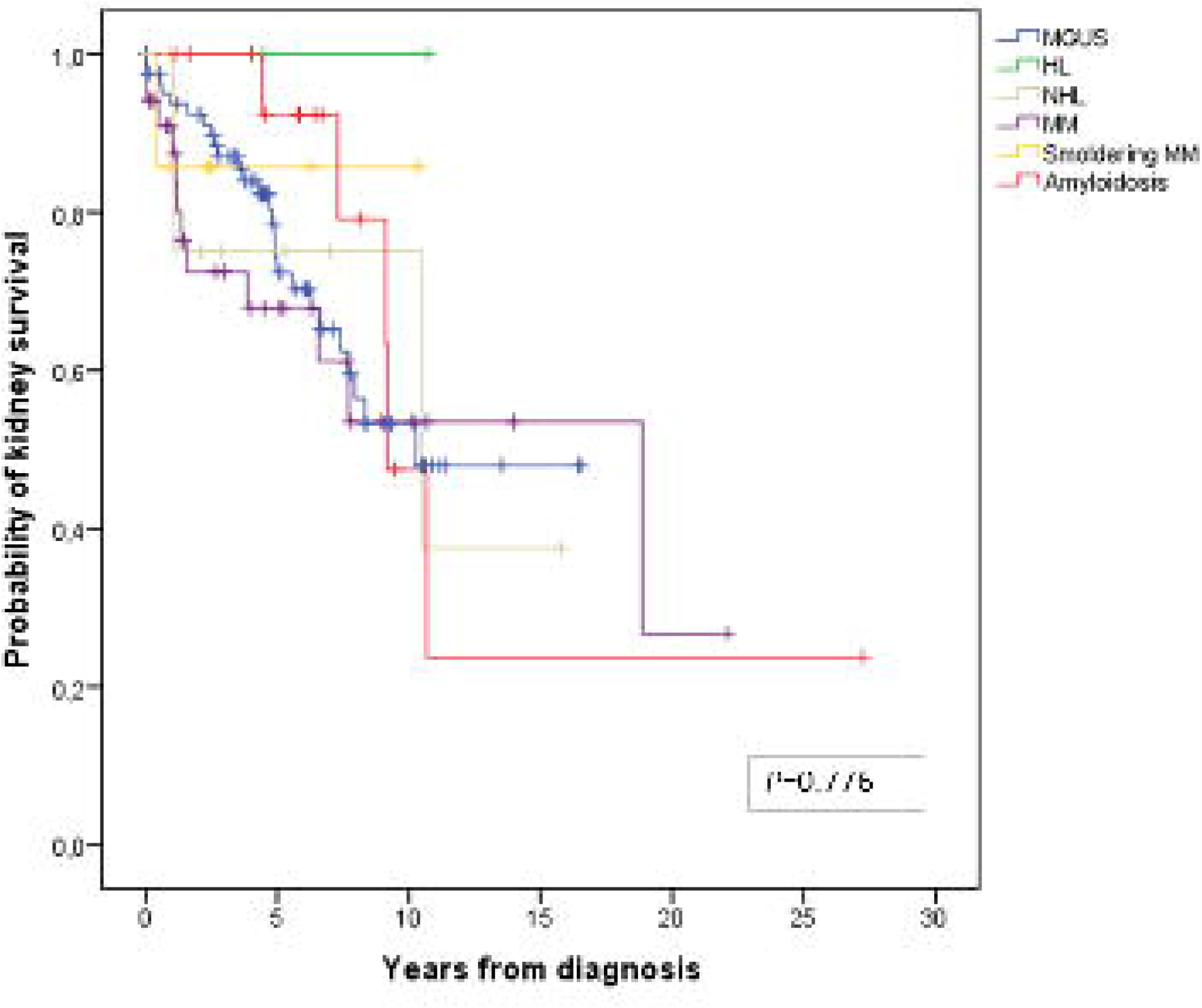
Death-censored kidney survival (CKD 5 or dialysis) according to the diagnosis of monoclonal gammopathy

## Discussion

The recent literature has placed great emphasis on the pathogenic role of monoclonal gammopathy as a potential cause of kidney disease. Our study showed that monoclonal gammopathy was a frequent diagnosis (13%) in patients with renal impairment who underwent kidney biopsy. Monoclonal gammopathy occurred predominantly in subjects aged more than 50 years with a peak over 70 years. In our cohort of patients, we diagnosed MGUS, SMM, NHL, LH, MM and AL amyloidosis. MGUS was the most common disorder. It accounted for more than half (52.8%) of all monoclonal gammopathies. Nevertheless, the prevalence rate of malignant lymphoproliferative disorders was surprisingly high in our cohort of patients, as a result MM, HL and NHL accounted for 42.8% of all gammopathies.

Besides the malignancy of these disorders, the nephrotoxicity of M-protein should be considered when evaluating monoclonal gammopathy. M-protein, even if it is produced by an indolent clone with a low propensity to progress toward uncontrolled proliferation, may be extremely harmful to renal parenchyma. Etiological mechanisms of M-protein nephrotoxicity are strictly dependent on the idiosyncratic properties of the secreted paraprotein. Deposition of M-protein[18] and activation of complement[19] are the leading pathological processes underlying the onset of monoclonal gammopathy-associated renal lesions[2]. According to the recent definition of MGRS[20], renal lesions due to the interplay with circulating M-protein were detected in 6.5% and 14.1% of MGUS and smoldering MM patients, respectively. Histological lesions compatible with MGRS included light chain deposition diseases and proliferative glomerulonephritis with M-immunoglobulin deposits. While light chain deposition disease is known to be associated with the deposition of circulation of M-protein[21], little is known on the role of M-protein in promoting membranoproliferative glomerulonephritis [22]. This latter histologic patter has been frequently encountered in patients with MGUS, but it is not infrequent in chronic lymphocytic leukemia, lymphomas and MM[23]. Deposition of secreted monotypic immunoglobulin protein along the capillary walls and the activation of the complement system (both classical and alternative pathway) are believed to be the main triggers for the development of the membranoproliferative pattern of glomerular damage.[22]

It is worth noting that the histological detection of kidney lesions with membranoproliferative glomerulonephritis patter is not sufficient to meet the diagnosis of MGRS. The identification of the restricted circulating immunoglobulin in renal parenchyma by immunofluorescence (or immunoperoxidase) and transmission electron microscopy is a practical and effective way to demonstrate direct M-protein nephrotoxicity[20]. Although the membranoproliferative glomerulonephritis patter was the predominant histopathological finding in MGUS and SMM patients, we found a few cases with Ig-restriction, corresponding to about one-tenth of all patients with membranoproliferative glomerulonephritis.

Among all lymphoproliferative disorders, MM was significantly associated with direct kidney injury (P=<0.0001). The etiological mechanism underlying kidney dysfunction was the production of a great amount of M-protein directly involved in the pathogenesis of myeloma-associated kidney disease. The majority of the patients with MM (81.8%) was admitted with AKI requiring renal replacement treatment in about a third of cases. In line with previous native renal biopsy studies[24,25], cast nephropathy was the most prevalent histopathological finding (68.1%), and as expected, it was also significantly associated with severe renal impairment. Interestingly, tubulointerstitial nephritis, a rare renal manifestation of MM, was found to be 9% of all cases[26].

Lymphoma is can be associated with kidney involvement presenting with a wide spectrum of manifestations. Lymphocytic infiltration of the parenchyma is the most prevalent finding in the largest case series of autopsies[27]. Further kidney manifestations rely on several distinct malignancy-related mechanisms and include minimal change disease, amyloidosis, membranoproliferative glomerulonephritis, immunotactoid glomerulopathy and M-protein deposition disease. In the setting of HL, presentation of kidney involvement in our series was mild proteinuria with normal renal function, but the limited number of cases does not allow us to generalize this data. On the other hand, renal function was extremely variable in NHL patients, ranging from normal renal function to acute kidney failure. Glomerulonephritis with membranoproliferative-like patterns and M-immunoglobulin deposition disease were the most common histological findings in this group of patients. Similar to the literature, glomerular involvement with membranoproliferative glomerulonephritis[28,28,29] and M-protein deposition disease[30–32] was common in patients with NHL.[33,34]

AL amyloidosis represented only a small percentage (8.9%) of all monoclonal gammopathies. AL amyloidosis was characterized by the deposition of light-chain deposition in the renal parenchyma of all renal biopsy specimens[35]. Our results confirmed the high prevalence of λ light chain isotype. AL amyloidosis manifested with a nephrotic syndrome characterized by a significantly higher level of proteinuria than other gammopathies. Renal function was not severely impaired and showed only a slight increase in sCr.

At the end of the follow-up, the evolution of renal function was extremely heterogeneous. The rate of ESRD or dialysis ranged from 0% (HL) to 38.2% (MGUS) without statistically significant differences among the groups of patients. In particular, renal outcome of MM patients was less dramatic than the initial stage of the disease. Recovery of renal function occurred in many of them, and the prevalence of ESRD or dialysis did not increase after 4.4±5 years of follow-up. Seven subjects with MGRS had a different renal prognosis at the end of the observation period, indeed in only two cases, CKD progressed to renal failure.

Survival of patients with malignant gammopathies was poorer than patients with a premalignant-clone. However, multiple factors (underlying kidney disease, disease-specific therapies, and supportive care) may have influenced the outcomes of these patients. In particular, the prognosis of patients with malignant disorders is changed in the past 10-15 years with the administration of promising therapeutic strategies such as proteasome inhibitor bortezomib, monoclonal antibodies, and the immunomodulatory drugs such as thalidomide and lenalidomide[36–38].

In clinical practice, the workflow process for the assessment of monoclonal gammopathy-associated renal lesion is based on the identification of the hematological disorder and underlying nephropathy. Once M-protein has been identified and characterized, exclusion of a malignant disorder should remain a high priority among nephrologists, as the outcome of the patient is associated with a poor prognosis if left untreated. Diagnostic tests such as flow cytometry, bone marrow biopsy, radiological examinations should have a low threshold if there is a high suspicion for lymphoproliferative disease. Evaluation of renal function trajectory and urinary abnormalities is essential to assess renal function. Kidney biopsy has a crucial role in the diagnosis of the underlying hematological disorder and renal injuries driven by M-protein. Lastly, kidney biopsy carries important therapeutic and prognostic implications in subjects with MGRS, as this condition is associated with a concerning poor renal outcome and with a high rate of recurrence after renal transplantation and appropriate chemiotherapy[39].

The main limitations of the study are the retrospective analysis and the small sample size of certain groups of patients with rare monoclonal gammopathies (especially HL and smoldering MM) that do not allow to generalize our results. The not routine use of electron microscopy has potentially led to the underestimation of some cases of MGRS and point out an unintentional bias frequently present in the current literature. The data collected over 17 years underlines the difficulty to categorize some kidney biopsies reporting a diagnosis of “membranoproliferative glomerulonephritis”. This term now refers to a histological pattern of glomerular lesions rather to a diagnosis of kidney disease. To avoid misclassification, we classified all “membranoproliferative glomerulonephritis” with the term “kidney injury with membranoproliferative pattern”.

## In conclusion

Lymphoproliferative disorders secreting M-protein carry a different potential for kidney injury. MGUS is the most frequent monoclonal gammopathy among patients who undergo kidney biopsy. Although MGUS has a low propensity to progress toward malignant disease, it is related to the development of specific renal lesions. MM is significantly associated with renal impairment and commonly manifesting with severe impairment of renal function. Patients diagnosed with AL amyloidosis has a higher level of proteinuria compared to the other monoclonal gammopathies. Careful evaluation is mandatory to identify malignant disorders and MGRS because both conditions require specific chemotherapy treatment and have a different prognosis compared to patients with pre-malignant monoclonal gammopathies.

## Data Availability

Available upon valid request

## Declarations

### Compliance with Ethical Standards

The study has been conducted ethically in accordance with the World Medical Association Declaration of Helsinki

### Ethics approval

The study has been approved by the Ethical Committee of Emilia Romagna

### Data available statement

The data underlying this article will be shared on a reasonable request to the corresponding author.

